# TMPRSS2-Coagulation Nexus: A Novel Molecular Link Revealed by Pairwise Correlation Analysis Following AstraZeneca (ChAdOx1 nCoV-19) Vaccination in a Nigerian Cohort

**DOI:** 10.64898/2026.06.20.26356158

**Authors:** Erens Spiff Ekprikpo, Z.A. Jeremiah, Stella U. Ken-Ezihuo, Beauty Eruchi Echonwere-Uwikor

## Abstract

**Background:** While haematological and coagulation changes following AstraZeneca vaccination have been described, the molecular mechanisms linking TMPRSS2 expression to coagulation remain underexplored, particularly in African populations.

**Methods:** In this case-control study, 102 adults (51 vaccinated with AstraZeneca ≥6 months prior, 51 unvaccinated controls) aged 18–65 years in Port Harcourt, Nigeria, were evaluated. Full blood count (Sysmex XN-1000), PT/aPTT (Erba Mannheim), RNA concentration, and qRT-PCR for ACE2/TMPRSS2 (normalized to GAPDH) were performed. Pearson correlations and t-tests were conducted (SPSS v26, p<0.05).

**Results:** Because primary between-group comparisons from this cohort have been reported previously, the present analysis focused on correlation patterns. A statistically significant inverse correlation was observed between TMPRSS2 Ct values and activated partial thromboplastin time (aPTT) among vaccinated participants (r = −0.325, p = 0.0202), whereas no corresponding association was detected in unvaccinated controls. Sex-specific differences in TMPRSS2 expression were also observed. No participant reported severe thrombotic or haemorrhagic complications at the time of recruitment.

**Conclusion:** This secondary analysis identified a statistically significant inverse correlation between TMPRSS2 Ct values and aPTT among AstraZeneca-vaccinated individuals. Although causality cannot be inferred, the findings suggest a potential relationship between TMPRSS2 expression and haemostatic pathways in the post-vaccination setting. Larger longitudinal studies are required to validate the observation and clarify its biological significance.

## Introduction

The AstraZeneca COVID-19 vaccine (ChAdOx1 nCoV-19) has been instrumental in global vaccination efforts, including in Nigeria.^1^ Clinical trials demonstrated that the vaccine provides substantial protection against symptomatic COVID-19 and severe disease while maintaining an acceptable safety profile across diverse populations.^2^ The World Health Organization (WHO) has identified the vaccine as a key component of global vaccination strategies, particularly in low- and middle-income countries where accessibility and storage advantages support widespread deployment.^3^

Previous studies from this cohort documented haematological alterations following AstraZeneca vaccination, including increased packed cell volume (PCV), elevated white blood cell count (WBC), and reduced platelet counts.^4^ A subsequent investigation also demonstrated changes in coagulation parameters and differential expression of ACE2 and TMPRSS2 genes among vaccinated individuals.^5^ Similar vaccine-associated haematological and immunological responses have been reported in other populations and are generally considered manifestations of immune activation following vaccination.^6^

Although the AstraZeneca vaccine has demonstrated a favourable overall safety profile, rare coagulation-related adverse events, including vaccine-induced immune thrombotic thrombocytopenia (VITT), have generated considerable scientific interest regarding the molecular mechanisms linking vaccination and haemostatic pathways.^7^ Current evidence has largely focused on platelet factor 4 (PF4)-mediated immune responses; however, other molecular mediators that may influence coagulation after vaccination remain insufficiently characterized.

TMPRSS2, a transmembrane serine protease, facilitates SARS-CoV-2 entry through proteolytic activation of the viral spike protein and subsequent ACE2-mediated membrane fusion.^8,15^ Beyond its established role in viral infectivity, TMPRSS2 belongs to a broader family of serine proteases involved in proteolytic regulation that may influence inflammatory and haemostatic pathways.^9^ Previous studies have suggested important interactions between inflammatory responses, endothelial activation, and coagulation dysregulation during SARS-CoV-2 infection and related immune processes.^10^

Despite increasing interest in TMPRSS2 biology, little is known regarding its potential relationship with coagulation parameters following COVID-19 vaccination, particularly in African populations. Furthermore, while previous studies from this cohort documented haematological alterations, coagulation changes, and differential ACE2/TMPRSS2 expression following AstraZeneca vaccination, the relationships among these variables have not been explored.^4,5^ The primary between-group comparisons of haematological, coagulation, and gene-expression parameters from this cohort have therefore been reported previously.^4,5^ The present study represents a secondary analysis focused specifically on pairwise molecular-haematological and coagulation correlations. Therefore, this study examined pairwise correlations between TMPRSS2 expression, coagulation indices, and haematological parameters among vaccinated and unvaccinated adults in Port Harcourt, Nigeria, to explore a potential TMPRSS2-coagulation nexus in the post-vaccination setting.

## Materials and Methods

### Study Design

This was a prospective case-control study conducted between January and December 2024 at the Rivers State University Teaching Hospital (RSUTH) and collaborating molecular laboratories in Port Harcourt, Rivers State, Nigeria. The study compared molecular (ACE2 and TMPRSS2 gene expression), coagulation (PT and aPTT), and haematological parameters between individuals who had received the AstraZeneca (ChAdOx1 nCoV-19) COVID-19 vaccine and age- and sex-matched unvaccinated controls.

### Ethical Approval

The study protocol was reviewed and approved by the Research Ethics Committee of the Rivers State Hospital Management Board (Approval No: RSHMB/RSHREC/2024/113). All procedures were performed in accordance with the Declaration of Helsinki. Written informed consent was obtained from every participant after a thorough explanation of the study objectives, procedures, potential risks, and benefits. Participation was voluntary, and participants could withdraw at any time without repercussions.

## Participants

### Inclusion Criteria

1. Adults aged 18–65 years.
2. Permanent residents of Port Harcourt, Rivers State, Nigeria.
3. Completion of the primary AstraZeneca COVID-19 vaccination series at least 6 months prior to enrolment (vaccinated group).
4. Apparently healthy unvaccinated individuals (control group).
5. Willing and able to provide informed consent.

### Exclusion Criteria

1. Age below 18 or above 65 years.
2. Refusal to provide written informed consent.
3. History of severe allergic reaction to any vaccine component.
4. Known contraindicating medical conditions, including autoimmune diseases, bleeding or thrombotic disorders, pregnancy or breastfeeding, acute or chronic infections requiring treatment, or current use of immunosuppressive drugs or anticoagulants.
5. Receipt of any COVID-19 vaccine other than AstraZeneca.

#### Sample Size

The required sample size was determined using G*Power software (version 3.1.9.4). Assuming a medium effect size (Cohen’s d = 0.5), a significance level of α = 0.05, and statistical power of 80%, a total of 102 participants (51 per group) was calculated and successfully recruited.

#### Sample Collection and Processing

Venous blood samples (10 mL) were collected from each participant under strict aseptic conditions. Blood was drawn into:

1. K_2_EDTA tubes for full blood count (FBC) and RNA extraction
2. 3.2% trisodium citrate tubes for coagulation assays

Coagulation samples were processed immediately to obtain platelet-poor plasma. EDTA samples for molecular studies were centrifuged, and aliquots were stored at –80°C until analysis to preserve RNA integrity.

### Laboratory Analyses

#### Full Blood Count (FBC)

Haematological parameters including packed cell volume (PCV), white blood cell count (WBC) and differential, platelet count, mean platelet volume (MPV), platelet distribution width (PDW), plateletcrit (PCT), and platelet-large cell ratio (P-LCR) were analysed using the Sysmex XN-1000 Automated Haematology Analyzer (Sysmex Corporation, Kobe, Japan) according to the manufacturer’s instructions.

#### Coagulation Studies

Prothrombin time (PT), activated partial thromboplastin time (aPTT), and International Normalized Ratio (INR) were determined using the Erba Mannheim ECL-105 semi-automated coagulometer (Erba Mannheim, Germany) with commercial reagents following standard laboratory protocols.

#### Molecular Analysis

Total RNA was extracted from whole blood using the Zymo Quick-RNA Plus Isolation Kit (Zymo Research, Irvine, CA, USA) according to the manufacturer’s protocol. RNA quantity and purity were assessed using a NanoDrop spectrophotometer. Complementary DNA (cDNA) was synthesized, and quantitative real-time PCR (qRT-PCR) was performed to evaluate the expression levels of *ACE2* and *TMPRSS2* genes, normalized to the housekeeping gene *GAPDH*. Cycle threshold (Ct) values were recorded for subsequent statistical analysis.

#### Statistical Analysis

Data were analysed using IBM SPSS Statistics for Windows, version 26.0 (IBM Corp., Armonk, NY, USA). Continuous variables are presented as mean ± standard deviation (SD). The normality of data distribution was assessed using the Shapiro-Wilk test. Comparisons between vaccinated and unvaccinated groups were performed using independent samples t-tests. Pearson’s correlation coefficient (r) was used to explore pairwise relationships between molecular (ACE2 and TMPRSS2 Ct values), coagulation, and haematological parameters. A two-tailed p-value < 0.05 was considered statistically significant.

## Results

Demographic characteristics of the study participants are presented in Table 1. The vaccinated and unvaccinated groups were well-balanced with respect to age, sex, marital status, and BMI.

**Table 1:** Demographic Characteristics of Study Population.

|  | Total | Vaccinated (Test) | Unvaccinated (Control) |
| --- | --- | --- | --- |

| Characteristics | N (%) | n (%) | n (%) |
| --- | --- | --- | --- |
| **Sex** |  |  |  |
| Female | 45 (44.1) | 18 (35.3) | 27 (52.9) |
| Male | 57 (55.9) | 33 (64.7) | 24 (47.1) |
| **Age Group (years)** |  |  |  |
| <30 | 29 (28.4) | 5 (9.8) | 24 (47.1) |
| 30-44 | 51 (50.0) | 35 (68.6) | 16 (31.4) |
| 45+ | 22 (21.6) | 11 (21.6) | 11 (21.6) |
| **Mean ± SD** | 36.8 ± 8.7 | 38.8 ± 6.3 | 34.8 ± 10.3 |
| **Marital Status** |  |  |  |
| Single | 47 (46.1) | 20 (39.2) | 27 (52.9) |
| Married | 55 (53.9) | 31 (60.8) | 24 (47.1) |
| **BMI** |  |  |  |
| **Mean ± SD** | 26.4 ± 4.2 | 26.8 ± 4.1 | 26.0 ± 4.3 |

Because the primary haematological, coagulation, and gene-expression comparisons from this cohort have been reported previously,^4,5^ the present analysis focused on pairwise correlations among molecular, coagulation, and haematological parameters.

The primary novel finding of this study was a statistically significant inverse correlation between TMPRSS2 Ct values and activated partial thromboplastin time (aPTT) in vaccinated participants (r = −0.325, p = 0.0202), representing a weak-to-moderate association. No corresponding relationship was observed among unvaccinated controls, suggesting that this association may be specific to the post-vaccination state (Table 2).

**Table 2:** Selected Pairwise Correlation Analysis of Coagulation and Molecular Parameters.

| Variable | by Variable | Vaccinated (Test)<br>n=51 |  | Unvaccinated (Control)<br>N=51 |  |
| --- | --- | --- | --- | --- | --- |
|  |  | Correlation<br>n | P-Value | Correlation | P-Value |
| PT (Sec) | INR | 0.182 | 0.2010 | 0.028 | 0.8445 |
| APTT (sec) | INR | 0.056 | 0.6988 | -0.071 | 0.6209 |
| APTT (sec) | PT (Sec) | 0.083 | 0.5644 | -0.005 | 0.9697 |
| RNA Conc.<br>(ng/uL) | INR | 0.224 | 0.1143 | -0.178 | 0.2102 |
| RNA Conc.<br>(ng/uL) | PT (Sec) | -0.117 | 0.4129 | 0.133 | 0.3529 |
| RNA Conc.<br>(ng/uL) | APTT (sec) | -0.010 | 0.9471 | 0.019 | 0.8944 |
| GAPDH (CT) | INR | 0.096 | 0.5038 | -0.036 | 0.8035 |
| GAPDH (CT) | PT (Sec) | 0.127 | 0.3735 | 0.169 | 0.2360 |
| GAPDH (CT) | APTT (sec) | 0.077 | 0.5922 | -0.070 | 0.6250 |
| GAPDH (CT) | RNA Conc.<br>(ng/uL) | -0.172 | 0.2274 | -0.070 | 0.6279 |
| ACE2 (CT) | INR | -0.141 | 0.3237 | 0.059 | 0.6829 |
| ACE2 (CT) | PT (Sec) | 0.243 | 0.0853 | -0.002 | 0.9868 |
| ACE2 (CT) | APTT (sec) | 0.173 | 0.2260 | 0.006 | 0.9688 |
| ACE2 (CT) | RNA Conc.<br>(ng/uL) | -0.137 | 0.3373 | -0.235 | 0.0974 |
| ACE2 (CT) | GAPDH (CT) | 0.231 | 0.1028 | 0.337 | 0.0157* |
| TMPRSS2 (CT) | INR | -0.161 | 0.2576 | -0.097 | 0.4973 |
| TMPRSS2 (CT) | PT (Sec) | -0.045 | 0.7544 | 0.045 | 0.7540 |
|  |  | Correlation | P-Value | Correlation | P-Value |
| TMPRSS2 (CT) | APTT (sec) | -0.325 | 0.0202* | 0.054 | 0.7090 |
| TMPRSS2 (CT) | RNA Conc.<br>(ng/uL) | -0.093 | 0.5182 | 0.548 | <.0001**** |
| TMPRSS2 (CT) | GAPDH (CT) | 0.041 | 0.7744 | 0.163 | 0.2517 |
| TMPRSS2 (CT) | ACE2 (CT) | 0.130 | 0.3624 | 0.204 | 0.1505 |

\*\*Significance level: \*p < 0.05; \*\*p < 0.01; \*\*\*p < 0.001; \*\*\*\*p < 0.0001

**Table 3:**
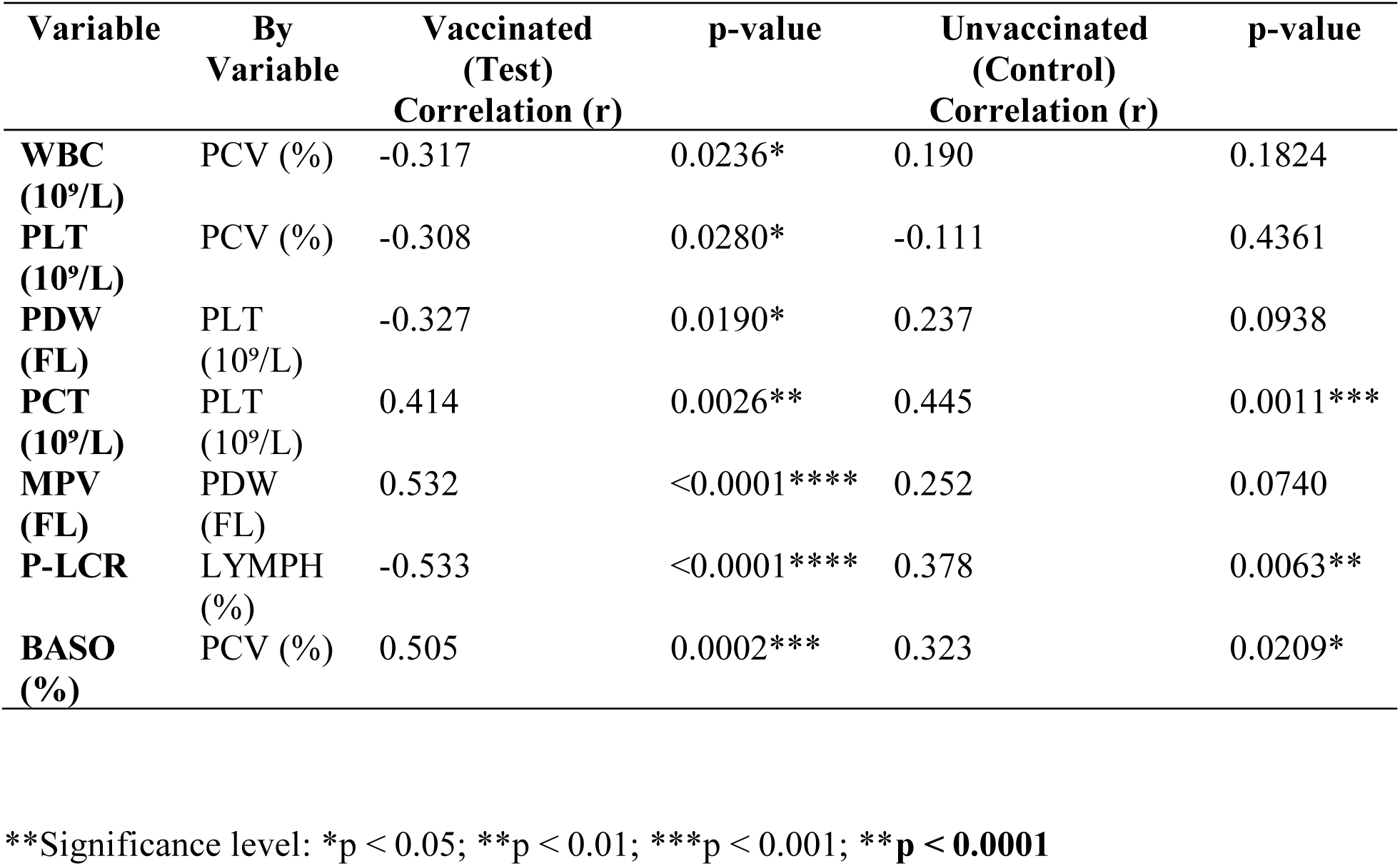
Selected Pairwise Correlation Analysis of Haematological Parameters in Vaccinated and Unvaccinated Subjects.

## Discussion

The present study identified a statistically significant inverse correlation between TMPRSS2 Ct values and activated partial thromboplastin time (aPTT) among vaccinated participants, whereas no such association was observed in unvaccinated controls. Because higher Ct values generally reflect lower gene expression, this finding suggests that alterations in TMPRSS2 expression may be associated with subtle changes in intrinsic coagulation pathway activity following vaccination. Although the observed correlation was modest and does not establish causality, it highlights a potential molecular-haemostatic interaction that warrants further investigation. TMPRSS2 belongs to the type II transmembrane serine protease family and has been studied primarily for its role in SARS-CoV-2 cellular entry; however, its broader biological functions may extend beyond viral infectivity to include interactions with inflammatory and proteolytic pathways relevant to haemostasis.^9^

Furthermore, increasing evidence suggests that inflammatory activation and coagulation are closely interconnected biological processes. Dysregulated protease activity has been implicated in endothelial dysfunction, thrombo-inflammatory responses, and haemostatic imbalance observed during SARS-CoV-2 infection.^13^ Although vaccine-induced immune responses differ fundamentally from active infection, overlapping molecular pathways may contribute to the subtle coagulation changes observed following vaccination. The absence of severe clinical events in the present study supports the overall safety profile of the AstraZeneca vaccine while highlighting the value of molecular monitoring approaches for understanding individual variability in vaccine responses.^14^

These results extend previous work from this cohort, which documented alterations in haematological parameters, coagulation profiles, and ACE2/TMPRSS2 expression following AstraZeneca vaccination.^4,5^ The present study moves beyond descriptive changes by identifying a statistically significant molecular-haematological relationship. The observed association may reflect interactions among immune activation, endothelial responses, and protease-mediated signalling pathways that occur following vaccination.

Importantly, no severe thrombotic or haemorrhagic events were observed among participants. This observation supports the established safety profile of the AstraZeneca vaccine while emphasizing that detectable molecular and coagulation changes do not necessarily translate into clinically significant adverse outcomes. Continued surveillance and larger longitudinal studies are warranted to clarify the long-term implications of TMPRSS2 modulation following vaccination.

Because multiple pairwise correlations were evaluated, the possibility of type I error should be considered when interpreting statistically significant associations. The observed relationship between TMPRSS2 Ct values and aPTT remained significant at the conventional α = 0.05 threshold; however, the exploratory nature of the analysis warrants cautious interpretation. Independent validation in larger cohorts is therefore required to confirm the robustness and reproducibility of this finding.

The findings should be interpreted within the context of certain limitations. The sample size was modest, and the cross-sectional design precludes causal inference. In addition, gene expression was assessed using Ct values as relative indicators of transcript abundance rather than absolute gene-expression quantification, which may limit biological interpretation. Future studies incorporating larger cohorts, longitudinal follow-up, cytokine profiling, and functional coagulation assays would provide deeper insight into the mechanistic pathways linking TMPRSS2 expression and haemostatic regulation.

## Conclusion

This secondary analysis of a previously characterized Nigerian cohort identified a statistically significant inverse correlation between TMPRSS2 Ct values and activated partial thromboplastin time (aPTT) among individuals vaccinated with the AstraZeneca (ChAdOx1 nCoV-19) vaccine. This association was not observed in unvaccinated controls, suggesting a potential relationship between TMPRSS2 expression and coagulation pathways in the post-vaccination setting. While the observed correlation was modest and should be interpreted cautiously given the exploratory nature of the analysis, it provides preliminary evidence supporting further investigation into the role of host proteases in vaccine-associated molecular and haemostatic responses. Overall, the findings are consistent with the established safety profile of the AstraZeneca vaccine and highlight the need for larger longitudinal studies to validate and clarify the biological significance of the observed association.

## Acknowledgements

The authors gratefully acknowledge the study participants for their time and valuable contribution. We are deeply thankful to our supervisors and co-authors -Prof. Z.A. Jeremiah, Dr. (Mrs.) Stella U. Ken-Ezihuo, and Dr. Beauty Eruchi Echonwere-Uwikor for their expert guidance, constructive criticism, and continuous support throughout the conduct of this research. Special appreciation is extended to the staff of the Rivers State University Teaching Hospital (RSUTH) Molecular Laboratory and MyAfro DNA Laboratory for their technical expertise and assistance during sample analysis.

## Funding

This research received no specific grant from any funding agency in the public, commercial, or not-for-profit sectors.

## Conflicts of Interest

The authors declare that they have no conflicts of interest.

## Data Availability Statement

The datasets generated and/or analysed during the current study are available from the corresponding author on reasonable request.

## Author Contributions

E.S. Ekprikpo conceived the study, collected data, performed statistical analyses, and drafted the manuscript. S.U. Ken-Ezihuo, B.E. Echonwere-Uwikor, and Z.A. Jeremiah supervised the study, reviewed the methodology, interpreted the findings, and critically revised the manuscript. All authors read and approved the final manuscript.

## Notes

### Competing Interest Statement

The authors have declared no competing interest.

### Author Declarations

The Ethics Committee of Rivers State Ministry of Health gave ethical approval for this work.

